# Evaluation and Clinical Validation of Monkeypox Virus Real-Time PCR Assays

**DOI:** 10.1101/2022.11.12.22282254

**Authors:** Margaret G. Mills, Kate B. Juergens, Jolene Gov, Carter McCormick, Reigran Sampoleo, Alisa Kachikis, John K. Amory, Ferric C. Fang, Ailyn C. Pérez-Osorio, Nicole A. P. Lieberman, Alexander L. Greninger

**Author notes:** Address correspondence to Alexander L. Greninger. Authors contributed equally to this work (Margaret G. Mills, Kate B. Juergens).

## Abstract

**BACKGROUND:** In spring of 2022, an outbreak of monkeypox spread worldwide. Here, we describe performance characteristics of monkeypox virus (MPXV)-specific and pan-orthopoxvirus qPCR assays for clinical use.

**METHODS:** We validated probe-based qPCR assays targeting MPXV-specific loci F3L and G2R (genes MPXVgp052/OPG065 and MPXVgp002 and gp190/OPG002, respectively) and a pan-orthopoxvirus assay targeting the E9L locus (MPXVgp057/OPG071). Clinical samples and synthetic controls were extracted using the Roche MP96 or Promega Maxwell 48 instrument. qPCR was performed on the AB7500 thermocycler. Synthetic control DNA and high concentration clinical samples were quantified by droplet PCR. Cross-reactivity was evaluated for camelpox and cowpox genomic DNA, vaccinia culture supernatant, and HSV- and VZV-positive clinical specimens. We also determined analytical performance of the F3L assay using dry swabs, Aptima vaginal and rectal swabs, nasopharyngeal, rectal, and oral swabs, cerebrospinal fluid, plasma, serum, whole blood, breastmilk, urine, saliva, and semen.

**RESULTS:** The MPXV-F3L assay is reproducible at a limit of detection (LoD) of 65.6 copies/mL of viral DNA in VTM/UTM, or 3.3 copies/PCR reaction. No cross-reactivity with herpesviruses or other poxviruses was observed. MPXV-F3L detects MPXV DNA in alternative specimen types, with an LoD ranging between 260-1000 copies/mL, or 5.7-10 copies/PCR reaction. In clinical swab VTM specimens, MPXV-F3L and MPXV-G2R assays outperformed OPXV-E9L by an average of 2.4 and 2.8 Cts, respectively. MPXV-G2R outperformed MPXV-F3L by 0.4 Cts, consistent with presence of two copies of G2R present in labile inverted terminal repeats (ITRs) of MPXV genome.

**CONCLUSIONS:** MPXV is readily detected by qPCR using three clinically validated assays.

## Introduction

Since its discovery in the 1970s, human cases of monkeypox (MPX) have rarely been reported outside of countries in western and central Africa where the causative agent, monkeypox virus (MPXV), is endemic (1). MPX is a zoonotic disease, and frequently causes isolated human infections through spillover events (2). Despite evidence of increased human-to-human transmission and warnings of global spread, the disease was largely neglected until its recent emergence in an outbreak beginning in spring of 2022 (3–5). By November2022, the epidemic had expanded to over 77,000 cases detected in more than 100 countries worldwide. In the United States, at time of writing, more than 28,000 cases have been reported, as well as several deaths (6). Worldwide, this outbreak of human monkeypox has disproportionately burdened men who have sex with men (7).

Individuals who contract MPXV may experience fever, lymphadenopathy, and a disseminated papillary rash that can lead to secondary bacterial infections (8, 9). Effects may include significant pain for weeks, as well as a wide range of complications including encephalitis, bronchopneumonia, and vision loss (10). Immunocompromised individuals, particularly those with AIDS, are at high risk of severe manifestations of MPXV, including death (11). Prophylactic vaccination with the JYNNEOS vaccinia virus vaccine and treatment with the antiviral tecovirimat (Tpoxx), both originally developed for variola (smallpox), are the major medical countermeasures available for MPXV. Because prevention of MPXV spread and effective treatment of MPXV infection inherently rely on rapid and early detection, development of clinical quantitative PCR (qPCR) assays is critical for patient care and to disrupt networks of transmission. In the context of a developing outbreak of an emerging virus that can be shed in multiple body fluids, evaluation of assay performance in many alternative specimen types is also critical to preparedness efforts (12, 13).

MPXV is a member of the orthopoxvirus (OPXV) family of double-stranded DNA viruses. Although they are large (100-300 kb), OPXV genomes are highly conserved across species, making MPXV-specific qPCR assay design a challenge. We evaluated two MPXV-specific qPCR assays that target the F3L and G2R loci, and compared the sensitivity, specificity, and accuracy of these assays to the CDC pan-orthpoxvirus assay OPXV-E9L (14–16). We also assessed performance of all three assays in the presence of alphaherpesviruses. We found MPXV-F3L and MPXV-G2R assays to have equivalent analytical sensitivity, with 100% sensitivity at the limit of detection (LoD) of 65.6 copies/mL. OPXV-E9L had a sensitivity of 95% at the same LoD. We demonstrated that the MPXV-F3L assay does not cross react with Camelpox virus (CMLV), Cowpox virus (CPXV) or Vaccinia virus (VACV) and detects MPXV DNA with adequate sensitivity in fourteen different specimen types.

## Materials and Methods

### Ethics and regulatory approvals

Use of excess clinical specimens was approved by the University of Washington Institutional Review Board with a consent waiver (STUDY00010205). Specifically, the use of excess breast milk and semen were approved as part of UW IRB-approved studies with informed consent (STUDY00000853, STUDY00001055, STUDY00008491).

### Primers and Probes

Primers and probes targeting the loci for MPXV-F3L (15), MPXV-G2R (14), and OPXV-E9L (16) were synthesized by ThermoFisher. Sequences used were as follows: F3L forward 5′-CATCTATTATAGCATCAGCATCAGA-3′ and reverse 5′-GATACTCCTCCTCGTTGGTCTAC -3′, probe 5′-FAM/TGTAGGCCGTGTATCAGCATCCATT/BHQ1-3′. G2R forward 5′-GGAAAATGTAAAGACAACGAATACAG-3′ and reverse 5′-GCTATCACATAATCTGGAAGCGTA-3′, probe 5′-FAM/AAGCCGTAATCTATGTTGTCTATCGTGTCC/BHQ1-3′. E9L forward 5′-TCAACTGAAAAGGCCATCTATGA-3′ and reverse 5′-GAGTATAGAGCACTATTTCTAAATCCCA-3′, probe 5′-VIC/CCATGCAATATACGTACAAGATAGTAGCCAAC/QSY-3′.

### MPXV DNA Absolute Quantification With ddPCR

Absolute quantities of standards used to create contrived MPXV-positive samples were determined using droplet digital PCR (ddPCR). Each ddPCR reaction was performed in quadruplicate and included 1.25μL water, 12.5μL ddPCR Mastermix for Probes (Bio-Rad), 1.25μL of 20x primer/probe mix (900 nM primers, 250 nM probe), and 10μL template DNA. Following droplet generation, amplification on the Bio-Rad C1000 Touch ddPCR system consisted of 10 min at 95°C; 40 cycles of 30 sec at 94°C, one min at 60°C; and 10 min at 98°C. Amplification was detected using QX200 Droplet Reader (Bio-Rad) and results analyzed with QuantaSoft Pro v1.0.596 software (Bio-Rad) (17).

### Nucleic Acid Extraction

All samples were handled in a biosafety cabinet prior to viral inactivation. DNA from breastmilk and semen specimens was extracted on the Promega Maxwell 48 with Maxwell RSC Viral Total Nucleic Acid Purification Kit. Breastmilk specimens were spun down for 10 minutes to separate the fat layer, and the liquid was used for extraction. Two hundred twenty microliters of sample were mixed with 200μL extraction mix (181.5μL lysis buffer, 18.2μL proteinase K, and 0.38μL EXOBS internal control). Samples were then incubated for 10 min at room temperature and 10 min at 56°C, then extracted following manufacturer protocols and eluted in 100 μL of Promega elution buffer.

DNA from all other specimen types was extracted on the MagNA Pure 96 with the DNA and Viral NA Small Volume Kit unless otherwise noted. Aptima collection tubes contain a reagent to inactivate MPXV, so 200μL from these tubes was extracted without additional lysis buffer. For lesion, nasopharyngeal, rectal, or oral swabs in viral transport media or universal transport media (VTM/UTM), cerebrospinal fluid (CSF), plasma, serum, whole blood, urine, or saliva, 100μL of sample was added to 100μL of buffer AL (Qiagen). Dry swabs were added to 1 mL VTM/UTM, incubated for 1 min, vortexed for 10 seconds, and treated as above. All samples were eluted in 100μL Roche elution buffer.

Select specimens were extracted on the MagNA Pure 96 with the DNA and Viral NA Large Volume Kit, with 250μL of sample added to 250μL of buffer AL (Qiagen), then eluted in 50μL Roche elution buffer.

### Qualitative PCR

All F3L and G2R qPCR reactions contained 2.71μL water, 11.95μL No-ROX QuantiTect master mix (Qiagen), 0.55μL ROX QuantiTect master mix (Qiagen), 0.1μL each of 100μM assay-specific forward and reverse primers, 0.05μL of 100μM assay-specific probe, 0.063μL EXOBS internal control primer/probe mix (18), 0.025μL of Uracil-N-Glycosylase (0.025 units, EpiCentre technologies), and 10μL of template DNA. All E9L qPCR reactions contained an additional 0.063μL water in place of EXOBS primer/probe mix. Amplification on the 7500 Real-Time PCR System (Applied Biosystems) consisted of 2 min at 50°C; 15 min at 95°C; and 45 cycles of 1 min at 94°C and 1 min at 60°C (17).

### Accuracy

Purified monkeypox DNA strain USA-2003 from BEI (NR-4928), and seven MPXV-positive remnant clinical specimens identified by Washington State Public Health Lab (WA PHL) using the OPXV-E9L primer/probe set, were used to establish accuracy (sensitivity) of the F3L and G2R assays. Remnant skin swab samples in VTM/UTM that tested negative using the OPXV-E9L assay were used to establish accuracy (specificity) of the F3L and G2R assays.

Synthetic MPXV DNA from ATCC (VR-3270SD) and high-titer MPXV-positive remnant clinical specimens identified by UWVL were used to spike into negative remnant clinical specimens, resulting in contrived positive specimens to confirm accuracy (sensitivity) of the F3L and G2R assays for all specimen types.

### Cross-Reactivity and Interfering Substances Analysis

To examine cross-reactivity of MPXV/OPXV assays with other orthopoxviruses, purified camelpox (CMLV) and cowpox (CPXV) viral genomic DNAs were obtained from BEI Resources (NR-50076 and NR-2641, respectively). Vaccinia virus culture supernatant (strain NYCHB, a kind gift from Dr. David Koelle) was extracted as for swab specimens. All samples were then assayed using the F3L, G2R, and E9L assays as described.

To test cross-reactivity of MPXV/OPXV assays with herpesviruses, 18 VZV-positive, 11 HSV1-positive, and four HSV2-positive skin swabs in VTM/UTM were selected from remnant clinical specimens. The HSV-positive specimens represented a range of HSV assay Cts (17.2-31.7). A subset of these VZV- and HSV-positive specimens were also spiked with VR-3270SD to create contrived double MPXV/VZV- or MPXV/HSV-positive specimens. All samples (MPXV-negative and -positive) were then assayed using the F3L, G2R, and E9L assays as described.

### Analytical Sensitivity and Limits of Detection

Limits of detection for F3L and G2R assays were determined for contrived positive specimens (synthetic DNA spiked into MPXV-negative remnant clinical specimens) in two stages. VR-3270SD was first serially diluted 10-fold in four pooled MPXV-negative skin swabs in VTM, with final concentrations of 3,300,000-33 copies/mL, and tested in 4 replicates per concentration. Twenty replicates were then each were tested at 330 and 33 copies/mL. Extractions and qPCR were performed as above.

Direct comparison of sensitivity/LoD for F3L, G2R, and E9L assays used a similar procedure, but with extractions utilizing the MP96 DNA/Viral NA Large Volume extraction kit: 250μL of each sample were mixed with 250μL buffer AL, extracted, and eluted in 50 μL elution buffer, representing a five-fold concentration. First, VR-3270SD was serially diluted in pooled MPXV-negative skin swabs in VTM, 10-fold dilutions from 33,000-33 copies/mL then finer dilutions between 330-33 copies/mL, and tested in 4 replicates per concentration. Second, 20 replicates each were tested at 330, 110, and 66 copies/mL. qPCR was performed as above with each assay.

### Analytical Sensitivity in Diverse Specimen Types

To test the specificity of the F3L assay in a variety of matrices, we gathered a total of 17-24 remnant clinical samples per specimen type. Nasopharyngeal, oral, and rectal swabs in UTM/VTM, CSF, plasma, serum, whole blood, urine, and saliva, came from UW Virology remnant clinical specimens. Semen and breastmilk were obtained from remnant clinical trial specimens from the UW Medical Center. Vaginal and rectal swabs in Aptima multitest tubes were obtained from Harborview Medical Center. All MPXV-negative specimens were extracted and PCR-amplified with F3L as described above.

To test the analytical sensitivity in these specimen types and establish a rough LoD in each, synthetic or viral MPXV DNA was spiked into n=20-24 samples per specimen type. VR-3270SD was spiked into CSF, plasma, serum, whole blood, nasopharyngeal swabs in VTM/UTM, breastmilk, and urine to a final concentration of 1,000 copies/mL. Once MPXV-positive specimens were available, a high-titer clinical specimen was spiked into specimens: into semen to a final concentration of 260 copies/mL; and into saliva, and oral and rectal swabs in VTM/UTM to a final concentration of 780 copies/mL. All contrived positive specimens were extracted and qPCR-amplified with F3L as described above.

Contrived positive dry swabs were made by diluting high-titer clinical specimen into pooled negative swab specimens in VTM/UTM, adding 50μL to each swab, and air-drying for 10 min. Once the swabs were dry, each was added to 1mL VTM, incubated for 10 min, and vortexed for 10 seconds. To establish an LoD for dry swabs, contrived positive samples were made and tested in two stages. First, the high-titer clinical specimen was serially diluted 10-fold to final concentrations of 5,300,000 - 530 copies/mL then finer dilutions between 5,300 - 530 copies/mL, and tested in 4 replicates per concentration. Second, 20 replicates each were tested at 1,600, 800, and 530 copies/mL (810, 400, and 270 copies/swab respectively). Dry swabs were processed, extracted, and qPCR-amplified with F3L as described above.

## Results

### Absolute quantification of synthetic control and high viral load clinical sample

Because MPXV genomic DNA and clinical samples were in short supply in spring 2022, we first validated synthetic DNA purchased from ATCC (VR-3270SD). This template contains primer and probe binding sites for several previously-developed assays, including F3L, G2R, and E9L, and has a copy number of 1×10^8^-1×10^9^ copies/mL DNA estimated by the manufacturer (14–16). We determined the precise number of copies of this standard using ten-fold serial dilutions tested in quadruplicate by ddPCR (Figure S1A-C). Measurements of VR-3270SD lot 70053297 yielded 3.28×10^8^ copies/mL DNA for F3L; measurements of VR-3270SD lot 70053666 resulted in equivalent concentrations for all three loci: 1.28×10^8^ copies/mL for MPXV-F3L, 1.30×10^8^ copies/mL for MPXV-G2R, and 1.29×10^8^ copies/mL for OPXV-E9L. In addition, a high viral load remnant clinical specimen (F3L Ct ∼16) used for downstream validation studies was quantified by ddPCR as 5.34×10^8^, 8.78×10^8^, and 4.78×10^8^ copies/mL for F3L, G2R, and E9L respectively. These data are consistent with equal copy numbers of loci present in the ATCC DNA standard and an extra copy of G2R present in MPXV genomes.

### MPXV-F3L and MPXV-G2R assays are highly specific for MPXV DNA

To determine specificity, we first tested 15 HSV-positive, 17 VZV-positive, and 52 known HSV/VZV-negative clinical remnant skin swabs in VTM/UTM using the CDC OPXV-E9L assay and confirmed that they were negative for MPXV. We verified that the F3L assay did not detect MPXV DNA in any of these specimens. The G2R assay similarly did not detect MPXV DNA in the negative or HSV-positive swabs, or in 16 out of 17 VZV-positive swabs; one VZV-positive specimen tested positive for G2R (Ct 39.8) and was negative on repeat (Table 1). Overall, the F3L assay had a negative percent agreement of 100% and the G2R assay had a negative percent agreement of 98.8% over 84 samples.

**Table 1:** MPXV-F3L, MPXV-G2R, and OPXV-E9L Sensitivity and Specificity.

| F3L | Known (Contrived) Positive | Known Negative |
| --- | --- | --- |
| Assay Positive | 31 | 0 |
| Assay Negative | 1 | 84 |

| G2R | Known (Contrived) Positive | Known Negative |
| --- | --- | --- |
| Assay Positive | 32 | 1 |
| Assay Negative | 0 | 83 |
| E9L | Known (Contrived) Positive | Known Negative |
| Assay Positive | 32 | 0 |
| Assay Negative | 0 | 84 |

*MPXV-F3L, MPXV-G2R, and OPXV-E9L assays are > 96.8% sensitive for MPXV DNA*

Camelpox virus (CMLV), Cowpox virus (CPXV) and Vaccinia virus (VACV) are the nearest phylogenetic relatives to MPXV based on DNA sequence similarity of conserved OPXV genes (16, 19). Accordingly, we tested VACV viral culture and purified CMLV and CPXV DNAs to assess cross-reactivity with MPXV-F3L and MPXV-G2R. The OPXV-E9L PCR assay served as a positive control for the other OPXV. VACV, CPXV, and CMLV were not detected by either the MPXV-F3L or MPXV-G2R assays, while the OPXV-E9L assay returned average Cts of 23.8, 25.8, and 19.8, respectively (Figure 1A).

**Figure 1.**
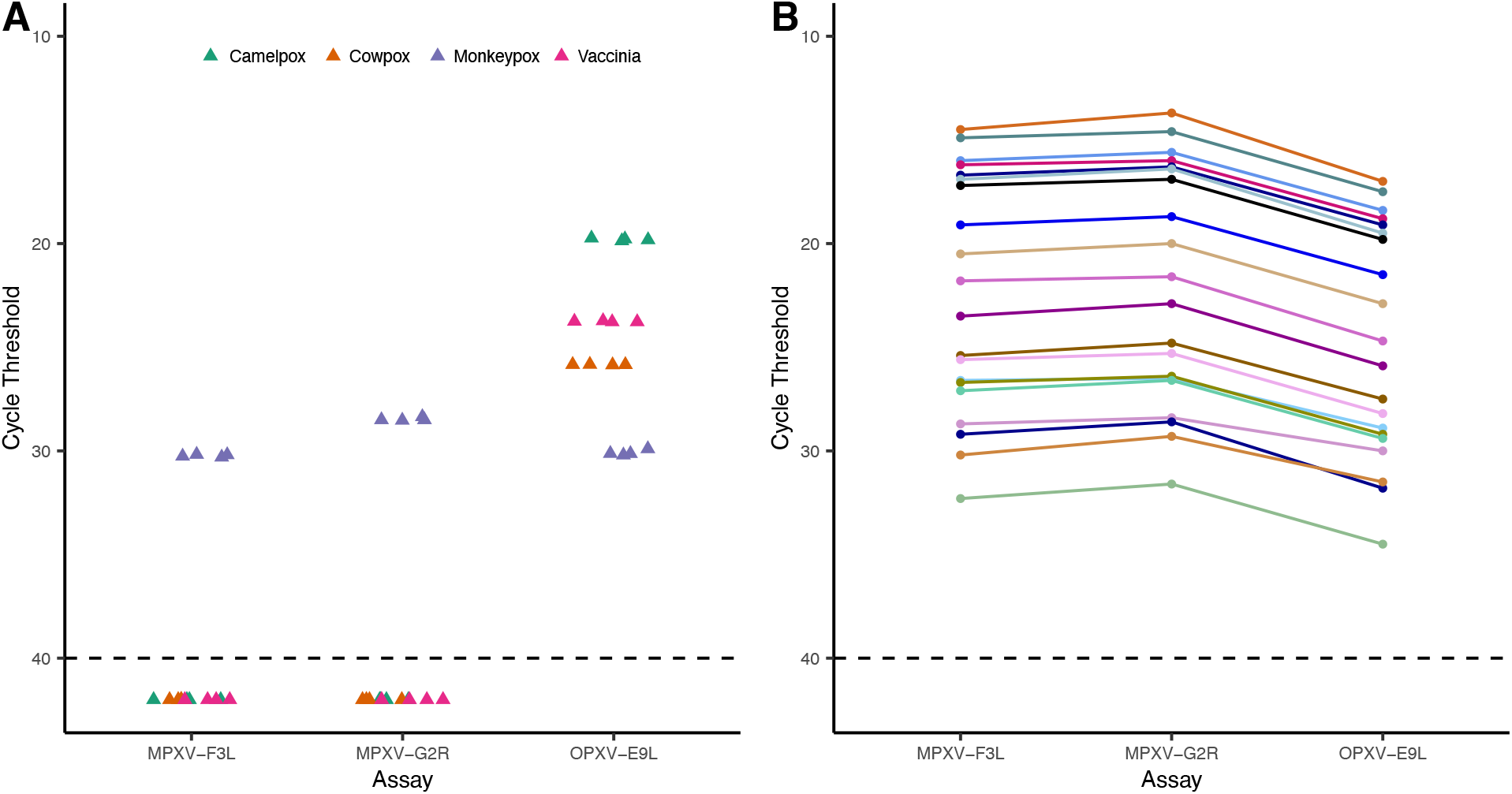
Specificity of MPXV-F3L and MPXV-G2R assays. (A) OPXV cross reactivity analysis of MPXV specific assays by testing MXPV, CPXV, CMLV, and VACV samples, with OPXV-E9L used as a positive control. Samples with no detectable MPXV are plotted at Ct of 42. (B). Specificity of all three assays with 20 clinical specimens initially positive by MPXV-F3L, followed by concomitant testing with MPXV-G2R and MPXV-E9L.

### MPXV-F3L, MPXV-G2R, and OPXV-E9L assays are > 96.8% sensitive for MPXV DNA

All three assays were tested for their ability to detect MPXV DNA using contrived positives consisting of a range of dilutions of VR-3270SD spiked into negative skin swabs in VTM (eight replicates at 330,000 copies/mL, 16 at 33,000 copies/mL, and eight at 3,300 copies/mL, based on ddPCR). G2R and E9L assays had 100% positive percent agreement, while a single replicate at 3,300 copies/mL was negative in the F3L assay (G2R Ct = 36.6), resulting in a positive percent agreement of 96.9%. Upon quadruplicate repeat testing, the failed sample had F3L Cts of 32.2, 33.4, 32.9, and 33.0. Overall percent agreement for the three assays were 99.1% for both G2R and F3L, and 100% for E9L.

### MPXV-F3L and MPXV-G2R assays detect MPXV viral DNA

Next, we confirmed that the F3L and G2R assays would accurately identify samples containing MPXV. Due to the short supply of MPXV-positive samples and genomic DNA early in the 2022 MPXV pandemic, we were only able to secure MPXV genomic DNA from BEI Resources, as well as seven MPXV-positive skin swabs in VTM from the WA State Public Health Laboratory (PHL). All known positive samples tested positive for MPXV by both F3L and G2R assays. Because of volume constraints, the PHL samples were diluted at least 1:40 prior to extraction at our lab, resulting in later Ct values for F3L and G2R assays than the Ct values for OPXV-E9L assay used by PHL, and complicating quantitative comparison of the assays. The qualitative interpretation was 100% concordant.

In order to quantitatively compare the ability of the three assays to detect MPXV DNA, we took 20 MPXV-positive clinical swab specimens in VTM (as detected by MPXV-F3L assay) and performed concomitant MPXV-G2R and OPXV-E9L testing on them. In every specimen, the MPXV-specific primers gave lower Ct values with averaging Ct differences of 2.4 (range, 1.3-2.9) and 2.8 (1.6-3.2) lower than OPXV-E9L for MPXV-F3L and MPXV-G2R assays, respectively. In addition, the MPXV-G2R gave a lower Ct value than MPXV-F3L in every specimen, averaging 0.4 Ct difference (0.1-0.9) (Figure 1B, Supplemental Table 1). This is consistent with two copies of G2R being present in the MPXV genome.

### MPXV-F3L, MPXV-G2R and OPXV-E9L assays are not affected by alphaherpesviruses

Co-infections of MPXV and HSV and VZV have been reported previously (7, 20). Therefore, we tested if the presence of HSV or VZV interfered with the detection of MPXV. We spiked VR-3270SD into HSV1-, HSV2-, or VZV-positive VTM specimens to a final concentration range of 330,000-3,300 MPXV copies/mL (n= 12 herpesvirus positive samples total). Contrived MPXV-positive controls with no herpesviruses (n=32) resulted in Cts with no significant difference from those MPXV/herpesviruses double positives for all three assays (Supplemental Figure 2, Supplemental Table 2, p>0.05, T test).

### MPXV-F3L, MPXV-G2R, and OPXV-E9L Assays Have Analytic Sensitivity of 3.3 copies per PCR reaction

We performed an initial LoD for MPXV-F3L, MPXV-G2R, and OPXV-E9L assays using contrived positive specimens containing VR-3270SD at a range of concentrations (33,000-33 copies/mL VTM) tested in quadruplicate. For both assays, all four replicates were detected at 33,000, 3,300, and 330 copies/mL, while only 2/4 and 3/4 of replicates amplified at 33 copies/mL with F3L and G2R, respectively. Throughout this range, linearity was excellent among positive samples (Supplemental Figure 3), with R^2^ values of 0.9990, 0.9986, and 0.9974 for F3L, G2R, and E9L assays, respectively.

Based on these results, we tested twenty additional replicates of contrived positive samples at both 330 copies/mL and 33 copies/mL with F3L and G2R assays. At 330 copies/mL, corresponding to 3.3 copies per PCR reaction, 19/20 and 20/20 replicates were detected for F3L and G2R respectively, with mean Ct values of 34.5 and 36.6 respectively. At 33 copies/mL, corresponding to only 0.3 copies per reaction, 4/20 and 3/20 replicates were detected for F3L and G2R respectively. Therefore, the limit of detection of both F3L and G2R assays is 330 copies/mL using standard extraction methods, or 3.3 copies per reaction.

To maximize our ability to distinguish between the three assays, an additional LoD study was performed using large volume extraction methods that provided a theoretical 5x concentration of DNA over the standard extraction methods (Supplemental Table 4). This study used both synthetic DNA (VR-3270SD, with equal concentration of all three targets) and high-titer clinical specimen diluted in negative clinical specimens. Similar to the previous LoD study, this was conducted in two stages. First, VR-3270SD was diluted 10-fold in MPXV-negative specimen VTM to final concentrations of 3,300,000-33 copies/mL as well as finer dilutions between 330 and 33 copies/mL, and was tested in quadruplicate. For all three assays, 4/4 replicates were detected at 3,300,000-330 and 110 copies/mL dilutions, while only 3/4, 3/4, or 1/4 replicate(s) amplified at 66 copies/mL with F3L, G2R, and E9L assays respectively. Second, 20 replicates each of both VR-3270SD and the high viral load clinical specimen were tested at 330, 110, and 66 copies/mL (copies of F3L for the clinical specimen). For VR-3270SD dilutions, 20/20 replicates amplified at 330 copies/mL with all assays; 19/20, 19/20, and 16/20 replicates amplified at 1.1e2 copies/mL with F3L, G2R, and E9L assays respectively; and 12/20, 11/20, and 13/20 replicates amplified at 66 copies/mL with F3L, G2R, and E9L assays respectively. For clinical specimen dilutions at 66 F3L copies/mL by ddPCR and a theoretical 3.3 copies/rxn, 20/20, 20/20, and 19/20 replicates amplified with F3L, G2R, and E9L assays respectively, with mean Ct values of 35.0, 33.9, and 36.6 respectively. Therefore, the LoD for each assay using the higher input volume was 110 copies/mL for the F3L and G2R assays and 330 copies/mL VTM for the OPXV-E9L assay based on the ATCC DNA material and 66 copies/mL VTM for each assay based on ddPCR-determined copy number of a viral positive.

### MPXV DNA is detected in diverse specimen types with an LoD of 1000 copies per mL or lower

Given the need for testing for MPXV in alternative specimen types in select clinical cases outside of diagnosing monkeypox infection, the UW Virology laboratory also validated the F3L assay in multiple other specimen types. Due to the rarity of many specimen types, we combined our sensitivity and specificity studies with LoD determination, using between 17-24 unique specimens of each type. Presumptive MPXV-negative cerebrospinal fluid, plasma, serum, urine, breastmilk, whole blood, nasopharyngeal/rectal/oral swabs in VTM/UTM, and vaginal/rectal swabs in Aptima tubes, were spiked to a concentration of 1000 copies/mL of VR-3270SD, followed by extraction and qPCR on both spiked and negative samples. The negative percent agreement for all specimen types was 100%. The positive percent agreement for Aptima vaginal and rectal swabs, nasopharyngeal, cerebrospinal fluid, plasma, breastmilk, and whole blood was 100%. The F3L assay was positive in 22/23 urine specimens and 21/22 serum specimens, for a positive percent agreement of 95.7% and 95.5% respectively (Figure 2, Supplemental Table 3). Therefore, the F3L LoD for these samples was determined to be 1000 copies/mL.

**Figure 2.**
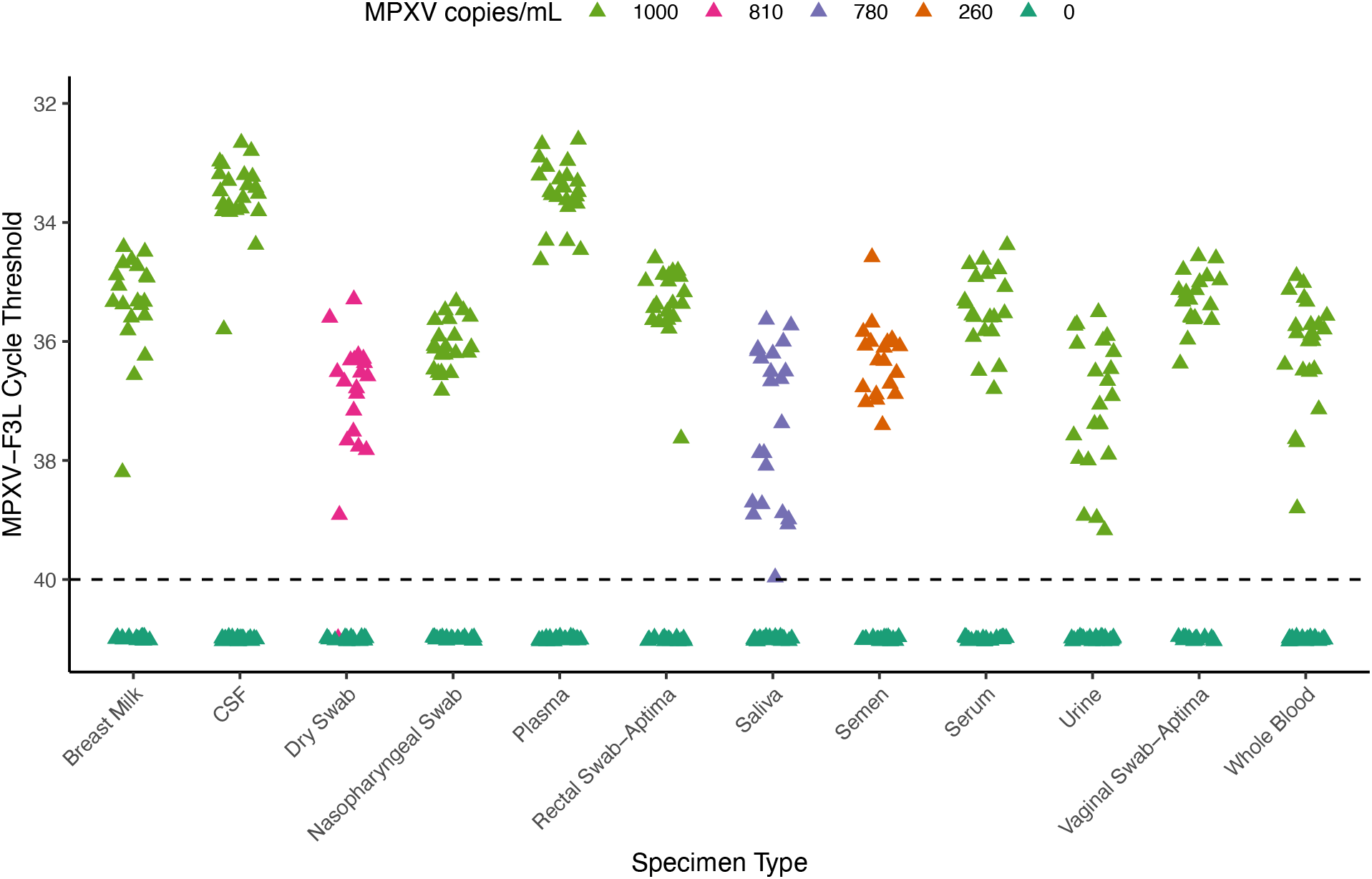
Sensitivity of MPXV-F3L With Diverse Specimens. MPXV-F3L Ct values of contrived MPXV positive breastmilk, CSF, dry swabs, nasopharyngeal swabs (NP), plasma, rectal swabs, saliva, semen, urine, vaginal swabs, and whole blood samples (n=17-24). Samples with no detectable MPXV are plotted at Ct of 41.

Once MPXV-positive specimens were available, and specifically for specimen types in which DNA degradation might be a concern (specifically dry swabs, semen, and saliva), we performed our validation using diluted MPXV clinical specimen rather than synthetic DNA. The high viral load clinical specimen was spiked into semen at 260 copies/mL, and into saliva and oral and rectal swabs at 780 copies/mL. Dry swabs were spiked with 810 copies per swab. Negative percent agreement of the F3L assay of semen, saliva, rectal, oral, and dry swabs was 100%. Positive percent agreement was 100% for semen, saliva, and rectal swabs, and 95% (19/20) for oral and dry swabs. Therefore, the F3L LoD for semen is 260 copies/mL (5.7 copies per PCR reaction), 780 copies/mL (7.8 copies per PCR reaction) for saliva, oral, and rectal swabs, and 810 copies/swab (8.1 copies per PCR reaction) for dry swabs.

## Discussion

Here, we evaluated the sensitivity and specificity of two MPXV-specific primer and probe sets in parallel with an orthopoxvirus assay during the ongoing 2022 MPXV pandemic. By extracting from up to 250 μL of specimen, we determined the LoD for MPXV-F3L, MPXV-G2R, and OPXV-E9L assays to be as low as 66 copies/mL, or 3.3 copies per PCR reaction. Our analytical sensitivity compares favorably with a recent evaluation of the PKamp Monkeypox Virus RT-PCR assay, which also uses the F3L gene as its target (21). Furthermore, all three assays did not cross react with HSV or VZV, demonstrating that both assays are highly specific to MPXV even in mixed samples with high HSV/VZV viral titer. We also evaluated the effect of sample matrix on the analytical sensitivity of the MPXV-F3L assay, finding that the assay was robust in fourteen different specimen types.

Although overall both MPXV-specific assays had very similar performance characteristics, F3L had a slightly lower Ct on average on synthetic positive control material and was present in a core genomic region, leading us to use it as our primary clinical assay for high-volume testing of swabs in VTM (17). In clinical samples, G2R detection was associated with slightly lower Ct values than F3L, likely due to the presence of two copies of the gene in the MPXV ITR regions. However, ITR regions of poxviruses are often subject to genomic rearrangements, especially as they spread to new hosts (22, 23). Reports of rare deletions in the G2R target that affected detection of MPXV DNA led the CDC to issue an advisory on September 2^nd^, 2022, urging caution in the interpretation of negative results returned from the G2R assay if clinical suspicion for MPXV was high (24). To date, no reports of deletions in F3L in MPXV have occurred. Altogether, we found very similar performance characteristics for the MPXV-F3L, MPXV-G2R, and OPXV-E9L assays for the detection of MPXV. Each assay was highly sensitive for MPXV DNA, with an LoD around 3 copies per reaction, near the limit of stochasticity. The F3L assay is specific for MPXV DNA, and does not cross-react with herpesviruses or other orthopoxviruses. This diagnostic will be an important defense for reducing transmission of MPXV during the ongoing outbreak.

## Data Availability

Majority data produced in the present work are contained in the manuscript. Additional data produced in the study are available upon request to the authors.

## Acknowledgements

The authors thank our study participants for their time and effort to donate specimens to the University of Washington Medical Center. AK is supported by NIH K23 AI153390-01.

## Notes

### Competing Interest Statement

ALG reports contract testing from Abbott, Cepheid, Novavax, Pfizer, Janssen and Hologic and research support from Gilead and Merck, outside of the described work.
Dr Kachikis reported serving as a research consultant for Pfizer and GlaxoSmithKline on maternal immunization-related projects in 2020 and as an unpaid consultant for GlaxoSmithKline in 2022 outside the submitted work. Dr Kachikis reported receiving grant support from Merck and Pfizer outside the submitted work.

### Funding Statement

This study did not receive any funding.

