## Supplemental Figures for "Evaluation and Clinical Validation of Monkeypox Virus Real-Time PCR Assays"

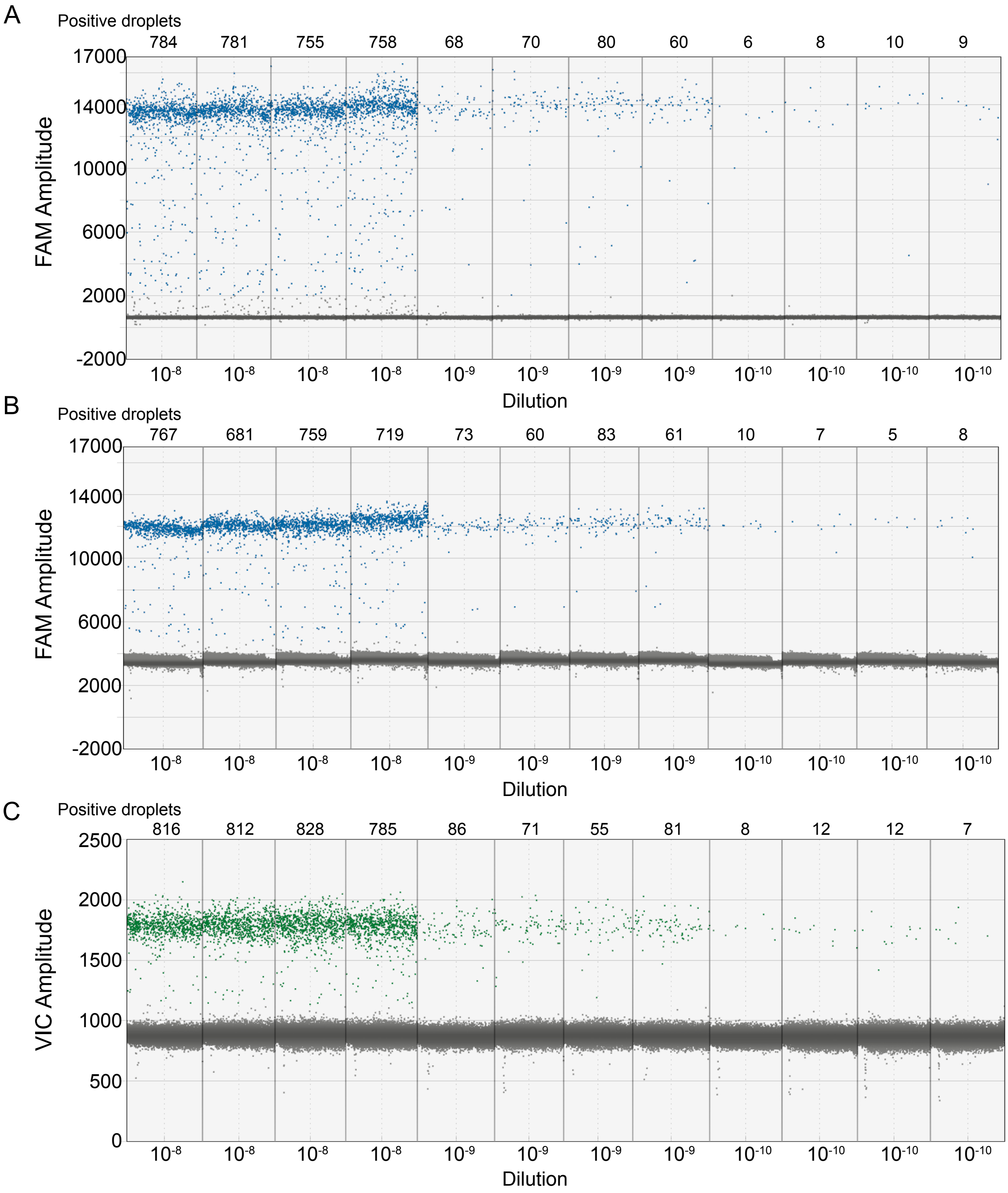


**Supplemental Figure 1.** Absolute quantification of synthetic MPXV DNA VR-3270SD with ddPCR. Threshold amplitude was set to 2031 for MPXV-F3L (A), 4763 for MPXV-G2R (B), and 1135 for OPXV-E9L assays (C). Serial dilutions of synthetic MPXV DNA are labeled across the x-axis. Absolute MPXV DNA-positive droplet counts are labeled above each assay.

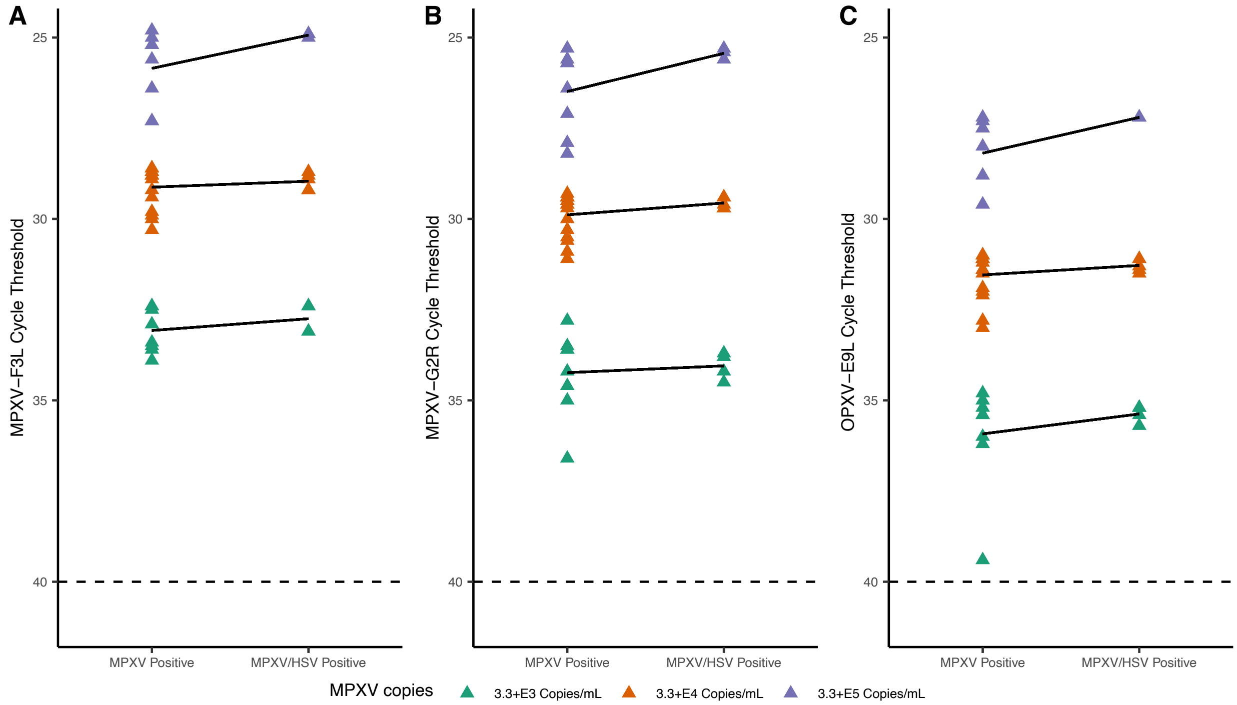


**Supplemental Figure 2.** Cross reactivity analysis of MPXV-F3L with serially diluted contrived MPXV-positive specimens with and without alphaherpesviruses.

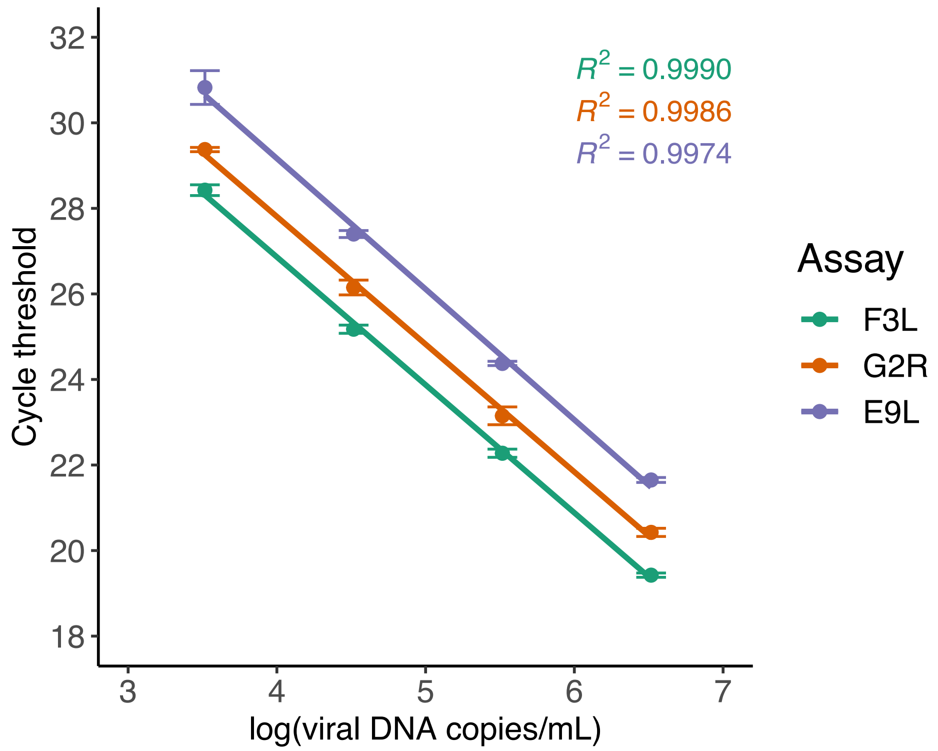


**Supplemental Figure 3.** Linearity of MPXV-F3L, MPXV-G2R, and OPXV-E9L assays using a panel of contrived MPXV positive specimens made with ATCC synthetic MPXV DNA.
